# Adherence to home-based videogame treatment for amblyopia in children and adults

**DOI:** 10.1101/2020.05.25.20113126

**Authors:** Tina Y. Gao, Joanna M. Black, Raiju J. Babu, William R. Bobier, Arijit Chakraborty, Shuan Dai, Cindy X. Guo, Robert F. Hess, Michelle Jenkins, Yannan Jiang, Lisa S. Kearns, Lionel Kowal, Carly S. Y. Lam, Peter C. K. Pang, Varsha Parag, Roberto Pieri, Rajkumar Nallour Raveendren, Jayshree South, Sandra Elfride Staffieri, Angela Wadham, Natalie Walker, Benjamin Thompson, on behalf of the BRAVO study team

## Abstract

**Objective:** Home-based videogame treatments are increasingly being used for various sensory conditions, including amblyopia (“lazy eye”), but adherence continues to limit success. To examine detailed behavioral patterns associated with home-based videogame treatment, we analyzed in detail the videogame adherence data from the <u>B</u>inocular <u>t</u>reatment of <u>a</u>mblyopia with <u>v</u>ide<u>o</u>games (BRAVO) clinical trial (ACTRN12613001004752).

**Methods:** Children (7-12 years), Teenagers (13-17 years) and Adults (≥18 years) with unilateral amblyopia were loaned iPod Touch devices with either an active treatment or placebo videogame and instructed to play for 1-2 hours/day for six weeks at home. Objectively-recorded adherence data from device software were used to analyze adherence patterns such as session length, daily distribution of gameplay, use of the pause function, and differences between age groups. Objectively-recorded adherence was also compared to subjectively-reported adherence from paper-based diaries.

**Results:** 105 of the 115 randomized participants completed six weeks of videogame training. Average adherence was 65% (SD 37%) of the minimum hours prescribed. Game training was generally performed in short sessions (mean 21.5, SD 11.2 minutes), mostly in the evening, with frequent pauses (median every 4.1 minutes, IQR 6.1). Children played in significantly shorter sessions and paused more frequently than older age groups (p<0.0001). Participants tended to over-report adherence in subjective diaries compared to objectively-recorded gameplay time.

**Conclusion:** Adherence to home-based videogame treatment was characterized by short sessions interspersed with frequent pauses, suggesting regular disengagement. This complicates dose-response calculations and may interfere with the effectiveness of treatments like binocular treatments for amblyopia, which require sustained visual stimulation.

**Clinical trial ID:** ACTRN12613001004752

## Introduction

Amblyopia, colloquially known as “lazy eye”, is a common neurodevelopmental visual condition that occurs in 1-3% of the population.[1] Most forms of amblyopia are characterized by reduced visual acuity in one eye and abnormal, unbalanced binocular vision, where the brain does not “use” the two eyes equally. Amblyopia is most commonly caused by childhood high anisometropia (large difference in refractive error between eyes), strabismus (misalignment of the eyes), or a combination of the two factors. In current standard clinical practice, amblyopia is treated in childhood with full-time wear of prescription glasses, followed by daily patching or atropine eye drops to penalize vision in the better-seeing eye for many months to years.[2] These long duration therapies are usually delivered at home by parents or caregivers, as office-based delivery for such prolonged treatment is costly and impractical. However, home-based treatments for amblyopia are often associated with poor treatment adherence.[3]

In the past decade, newer digital treatments targeting binocular vision have emerged, aided by improvements in display technologies like 3D monitors and virtual reality systems that enable separate images to be shown to each eye (dichoptic presentation). Binocular treatments rely on repeated exposure to visual stimuli that are biased in favor of the amblyopic eye and, theoretically, activate binocular neural circuits. Biasing of visual stimuli in favor of the amblyopic eye, a process referred to as *binocular-balancing*, can be achieved through altering image contrast[4], brightness[5], clarity[6], and/or spatial composition[7] independently for the two eyes. Repeated exposure is achieved through presenting these image manipulations within videogames or passive media such as movies. With many hours of exposure over periods of weeks or months, binocular treatments are hypothesized to “rebalance” the amblyopic visual system, producing improvements in visual function.[4]

Contrast-balanced binocular videogames are one particular type of binocular treatment which use reduced contrast images in the non-amblyopic eye and full contrast images in the amblyopic eye.[4, 8] The amount of contrast change is individually set for each amblyopic patient at the start of training, and the contrast difference between eyes is gradually reduced over the training period. Different game image components are seen by each eye so that visual information from both eyes must be combined to successfully play the game. Despite early promise in laboratory studies[8, 9] and home-based pilots[10, 11], recent larger-scale randomized clinical trials using home-based implementations of this type of videogame have found mixed results, ranging from greater efficacy than traditional patching therapy[11, 12], comparable efficacy to patching[13, 14], to no difference from placebo[15] or glasses wear[16]. Notably, several trials[13-16] reported a lack of dose-response relationship between visual gains and treatment adherence, which would suggest a lack of efficacy for binocular-balanced visual stimulation. However, these dose-response analyses use the cumulative game-playing time summed across the entire treatment period but do not account for whether binocular-balanced visual stimulation was received in hour-long blocks similar to the early successful laboratory studies[8, 9] or whether binocular stimulation was received in many short bursts distributed throughout the day. The latter pattern is more likely to occur in home-based videogame studies, particularly when portable devices are used for game implementation, as patients need to fit their daily treatment around other activities and often find it more convenient to perform treatment in shorter sessions. It is possible that frequent distractions or short session lengths reduce the effectiveness of binocular visual stimulation, thus reducing treatment effectiveness overall.

For practical reasons, future amblyopia therapies are likely to continue to be provided in unsupervised home settings. Thus, it is vital to understand the behavioral patterns associated with home-based videogame treatment in order to improve delivery methods and fully gauge treatment effectiveness. These adherence patterns are also useful to consider when designing other long-duration videogame treatments for disorders such as tinnitus[17] or traumatic brain injury[18].

To examine videogame treatment adherence patterns, we conducted *post hoc* analyses of data from participants who completed 6 weeks of at-home training in the <u>B</u>inocular <u>t</u>reatment for <u>a</u>mblyopia using <u>v</u>ide<u>o</u>games (BRAVO) clinical trial (ACTRN12613001004752). The BRAVO trial tested an active Tetris-based contrast-balanced videogame against a placebo videogame in children and adults with amblyopia, and found highly variable cumulative adherence, no dose-response association, and no significant differences in visual outcomes between active and placebo groups.[15] During the trial, we received anecdotal reports from participants (and parents of child participants) regarding disengagement, boredom, and multi-tasking while playing the game. This suggested that participants were perhaps not consistently attending to the visual stimuli, motivating the detailed adherence analyses we report in the current article. Specifically, we wanted to address the following questions: 1) What were the temporal patterns of game-play? 2) Was adherence pattern related to age? 3) Did adherence patterns differ between the active and placebo groups? And 4) Did objectively-recorded and self-reported adherence differ?

## Materials and Methods

The BRAVO study was an international placebo-controlled randomized clinical trial which compared a contrast-balanced falling-blocks (Tetris-like) videogame versus a placebo videogame for treatment of unilateral amblyopia.[15, 19] The trial was conducted at five study sites in four countries: New Zealand, Australia, Hong Kong, and Canada. Institutional ethical approval was obtained from the University of Auckland Human Participants Ethics Committee, the University of Waterloo Research Ethics Committee, the McGill University Health Centre, the Human Research and Ethics Committee of the Royal Victorian Eye and Ear Hospital, and the Human Subjects Ethics Subcommittee of the Hong Kong Polytechnic University. The trial adhered to the principles in the Declaration of Helsinki. Written informed consent was obtained for all adult participants and parents/guardians of younger participants, with either written or verbal assent from younger participants before enrolling in the study. Participants (and parents/guardians where relevant) were free to withdraw at any time, without needing to state a reason. The full trial protocol,[19] and main outcomes[15, 20] are described in previous publications.

After an appropriate optical correction only phase where needed[20], 115 eligible participants (age 7-55 years) with unilateral amblyopia associated with anisometropia and/or strabismus were randomized to either active (n=56) or placebo (n=59) videogame treatment with minimization stratification by age group. The three age groups were Children (7-12 years old), Teenagers (13-17 years old), and Adults (≥18 years old).

### Videogame treatment

Treatment games were implemented on 5^th^ generation Apple iPod Touch devices. Red-green anaglyphic glasses were used to produce dichoptic presentation. Both the active and placebo games were based on Tetris, a game where falling shapes are tessellated together to form complete rows of blocks. The active and placebo versions contained identical game levels, button controls, adjustable difficulty, and scoring mechanics, only differing in the type of visual stimuli presented. The active game[8, 10] presented blocks dichoptically (different blocks shown to each eye) with a contrast offset between the two eyes, requiring information from both eyes to be combined to successfully play. The placebo game presented identical, equal contrast elements to both eyes like a normal videogame, and could be played successfully even if participants did not use both eyes together.

All participants were instructed to play at home for 1-2 hours per day, every day, for 6 weeks while wearing the red-green glasses on top of any corrective glasses or contact lenses. Participants were free to split their videogame training into multiple sessions to fit around other activities, allowing us to observe their natural behavior during at-home videogame training.

### Objectively-recorded adherence

The game software continuously recorded a logfile, which contained detailed data such as when the game app was opened or closed, stimuli contrast, game scores, and in-game pauses. These data were extracted from the iPod devices and analyzed using custom programs written in MATLAB (2018a).

### Subjectively-reported adherence

All participants (and parents/guardians of children) were asked to record daily training times and high scores in a paper-based diary. We calculated the percentage concordance between self-reported and objectively-recorded adherence for each participant as follows:

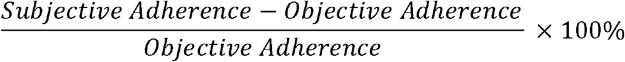

Positive percentage values indicated over-reporting of adherence in the subjective diary compared to the objective logfile, and negative values indicated under-reporting. Participants were not specifically asked to account for pauses when recording their treatment diary, so the “objective adherence” total used for this comparison was the total time that the game app was open, including both game-playing and paused time.

### Statistical analyses

Participants with missing logfile data, for example due to withdrawal from the study or refusal to play the treatment game, were excluded from all analyses. For the comparison between subjectively-reported and objectively-recorded treatment adherence, participants who did not return a diary were also excluded.

Two-way analyses of variance (ANOVA), with factors of game allocation (Active, Placebo) and age group (Child, Teenager, Adult) were used to investigate the following variables: cumulative time spent playing, cumulative time spent paused, average session length (length of time from opening the game app to closing the game app), frequency of pauses, average game scores, the number of days during the training period where the game was played (“training days proportion”), and treatment adherence on days where the game was played (“training days adherence”). The last two adherence variables were calculated based on methods described in Wallace, Stewart [3] to allow comparison with adherence to traditional occlusion therapy for amblyopia.

Weekday and weekend average daily adherence were analyzed using a two-way mixed ANOVA with a between-subjects factor of age group and a within-subjects factor comparing weekdays to weekend days. A Greenhouse-Geisser correction was applied to the within-subjects factor.

Analyses were performed in Matlab 2018a using the Statistics and Machine Learning Toolbox. Tukey-Kramer corrections were used for multiple comparisons.

## Results

### Overall adherence patterns

Out of 115 randomized participants, 10 (8.7%) participants did not complete 6 weeks of treatment[15], due to early withdrawal and/or refusal to play the videogame. Data from these 10 participants were excluded from the below adherence analyses.

The 105 participants who completed 6 weeks of treatment played on average only 65% (standard deviation [SD] 37%) or 27.5 (SD 15.7) hours of the prescribed minimum dose of 42 hours across 6 weeks (Table 1, Figure 1A). Individual cumulative adherence ranged widely from 1.8 hours (4.3% of the prescribed minimum) to 86.9 hours (207%, or about 2 hours per day).

**Table 1.** Videogame treatment adherence pattern summary.

|  |  | Children 7-12 years |  | Teenagers 13-17 years |  | Adults ≥18 years |  | Overall |  | Two-way ANOVA p-values |  |  |
| --- | --- | --- | --- | --- | --- | --- | --- | --- | --- | --- | --- | --- |
|  |  | Active | Placebo | Active | Placebo | Active | Placebo | Active | Placebo | Treatment Group | Age Group | Interaction |
| **Participants randomized** |  | **n=22** | **n=23** | **n=8** | **n=9** | **n=26** | **n=27** | **n=56** | **n=59** |  |  |  |
| **Completed 6 weeks of training** | **No. (%)** | **17(77)** | **23(100)** | **8(100)** | **9(100)** | **23(88)** | **25(93)** | **48(86)** | **57(97)** |  |  |  |
| Age (years) | Mean (SD) | 10.2(1.6) | 10.1(1.6) | 15.3(1.3) | 14.4(1.4) | 36.8(9.1) | 34.4(10.8) | 23.8(14.2) | 21.4(13.7) |  |  |  |
| Cumulative play time over 6 weeks (hours) | Mean (SD) | 24.8(18.4) | 23.9(14.3) | 23.9(17.0) | 31.3(14.4) | 28.7(14.4) | 31.3(16.9) | 26.5(16.1) | 28.3(15.6) | 0.37 | 0.26 | 0.66 |
| Proportion of minimum prescribed dose received (%) <sup>a</sup> | Mean (SD) | 59(44) | 57(34) | 57(40) | 75(34) | 68(34) | 75(40) | 63(38) | 67(37) |  |  |  |
| Cumulative pause time over 6 weeks (hours) | Mean (SD) | 1.2(0.9) | 2.2(3.7) | 0.6(0.5) | 0.8(0.6) | 0.8(1.3) | 0.6(0.7) | 0.9(1.1) | 1.3(2.5) | 0.41 | **0.04** | 0.40 |
| Training days proportion (%) | Mean (SD) | 71(31) | 77(25) | 67(33) | 80(24) | 79(22) | 80(25) | 74(27) | 79(24) | 0.23 | 0.58 | 0.71 |
| Average training day adherence (minutes played) | Mean (SD) | 42.5(20.6) | 38.6(17.5) | 43.6(18.9) | 51.5(15.0) | 47.1(16.8) | 51.1(18.5) | 44.9(18.3) | 46.1(18.4) | 0.50 | 0.09 | 0.45 |
| Average session length (minutes) | Mean (SD) | 13.6(6.2) | 15.8(8.4) | 25.8(16.3) | 24.9(10.4) | 25.6(12.8) | 25.9(8.3) | 21.4(12.8) | 21.6(9.8) | 0.64 | **<0.001** | 0.74 |
| Average playing time within a session (minutes) | Mean (SD) | 13.0(6.3) | 14.5(7.9) | 25.2(16.2) | 24.2(10.3) | 24.8(12.1) | 25.3(8.2) | 20.7(12.5) | 20.8(9.8) | 0.86 | **<0.001** | 0.90 |
| Average paused time within a session (minutes) | Mean (SD) | 0.7(0.3) | 1.2(1.8) | 0.6(0.4) | 0.7(0.7) | 0.8(1.7) | 0.5(0.7) | 0.7(1.2) | 0.9(1.3) | 0.57 | 0.45 | 0.83 |
| Average duration of play between pauses (minutes) | Mean (SD) | 2.4(1.6) | 2.4(2.0) | 8.6(9.3) | 8.1(6.7) | 8.9(7.6) | 12.8(12.1) | 6.5(7.1) | 7.9(9.7) | 0.50 | **<0.001** | 0.41 |
| Average game score (points) | Mean (SD) | 746(765) | 1279(815) | 2253(1107) | 2640(1647) | 2091(1238) | 3400(1263) | 1641(1245) | 2424(1518) | **0.003** | **<0.001** | 0.18 |
| Diary returned to research staff | No. (%) | 16(94) | 22(96) | 8(100) | 8(89) | 19(83) | 25(100) | 43(90) | 55(96) |  |  |  |
| Difference between diary and objective adherence, playing time only (%) <sup>b</sup> | Median (IQR) | 40(57) | 26(116) | 37(666) | 2(26) | 2(34) | 3(19) | 25(52) | 4(41) |  |  |  |
| Difference between diary and objective adherence, including paused time (%) <sup>b c</sup> | Median (IQR) | 31(57) | 12(103) | 30(655) | 1(25) | -1(33) | 3(16) | 16(51) | 3(30) |  |  |  |

a The minimum prescribed dose was 1 hour/day, or 42 hours over 6 weeks.
b Positive median values indicate participants over-reported playing time compared to the objective software recording, and negative values indicate under-reporting.

**Figure 1:**
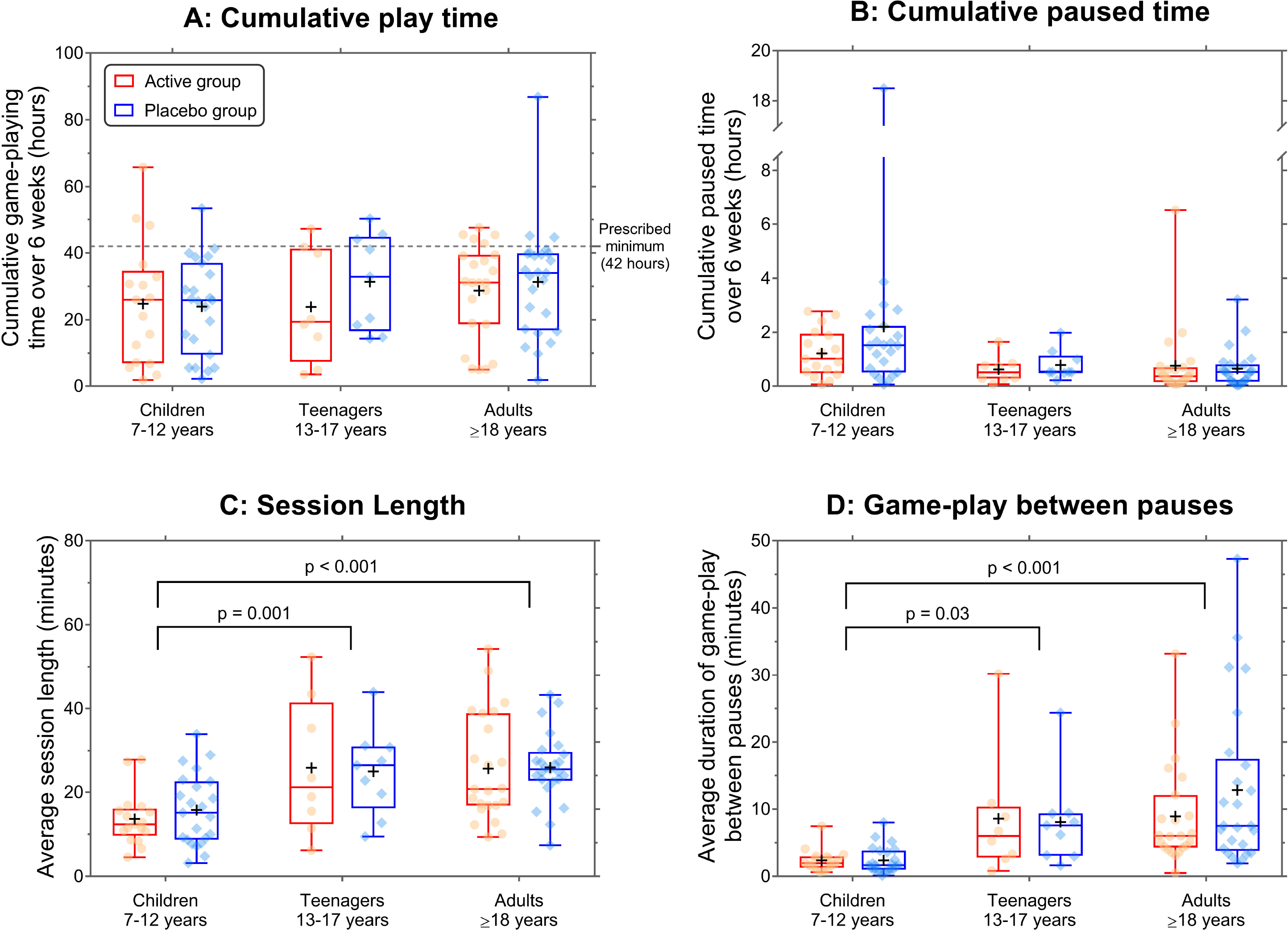
Videogame treatment adherence by age group and game allocation. A: Cumulative game-playing time over 6 weeks B: Cumulative game paused time over 6 weeks C: Average game session lengths D: Average game-playing time between pauses Box-plots indicate the median and quartiles, and whiskers extend to the full range of the data. Circle and diamond symbols indicate individual participant data. Black crosses indicate means for each group.

The average game session lasted 21.5 (SD 11.2, range 3.1 – 54) minutes. The distribution of session lengths (Figure 1C) suggests that most participants split their 1-2 hours per day of game-playing into multiple shorter sessions.

Within game-playing sessions, the game was paused on average every 4.1 minutes (median; interquartile range [IQR] 6.1; Figure 1D). There were 10 participants (including 8 Children) who averaged more than one pause per minute of game-play, consistently over the entire treatment period, which may indicate a severe lack of attention.

### Effects of treatment group and age groups

Compared to the Teenager and Adult groups, Children trained in shorter sessions, paused more frequently, and achieved lower average game scores (all p<0.04; Table 1, Figures 1C, 1D, S1). Active group participants also had lower average scores than Placebo group participants (Table 1, Figure S1). This is likely because the dichoptic nature of the active game made it more difficult to play than the placebo game, particularly at high speeds.

No significant main effects of game allocation or age group were found for cumulative game playing time, cumulative pause time, training days proportion, or training days adherence (all p>0.09; Table 1, Figures 1A, 1B, S2-S3). As there were no significant differences between the Active and Placebo groups apart from game scores, the two allocation groups were combined for all subsequent analyses relating to the temporal pattern of gameplay.

### Adherence patterns across 6 weeks

Detailed day-by-day adherence trends across the 6-week treatment period are shown in supplementary Figures S4-S6. In all three age groups, adherence peaked on the first full day of treatment (day 2 from randomization), followed by a gradual fall over the six-week period. This fall in adherence appeared to be mainly due to missed days of training (proportion trained) and, to a lesser extent, a fall in adherence on days where training was performed (training days adherence).

Average daily adherence did not significantly differ between weekdays and weekends (p=0.15) for any age group (Figure S7). Peak videogame play took place between 4-10pm for Children and between 6pm to midnight for Teenagers and Adults (Figure S8).

### Self-reported adherence

Ninety-eight (93%) of 105 participants returned their training diary. As shown in Figure 2, participants generally over-reported adherence compared to the objective logfile recording (median: 7.2% [IQR: 46.1%]). Just under half of the analyzed participants (47%, 46 out of 98) over-reported game-playing time by at least 10%, and within this, 29% (28 out of 98) over-reported by 30% or more. This over-reporting appeared to be most severe in the Children 7-12 years group (Table 1 and Figure 2).

**Figure 2:**
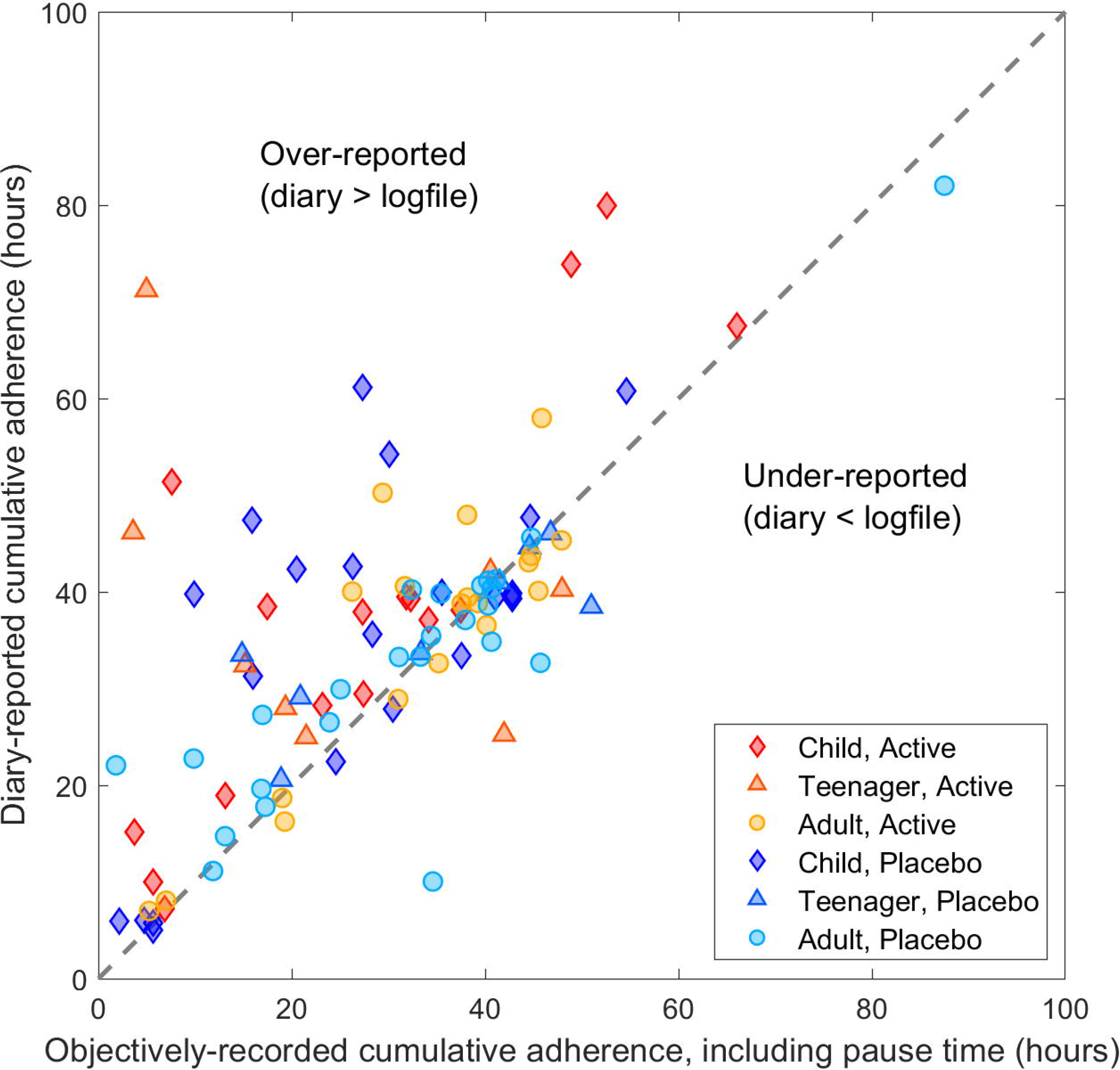
Concordance between subjectively-reported versus objectively-recorded cumulative adherence.

## Discussion

Adherence to home-based videogame treatment in the BRAVO trial was substantially less than prescribed and highly variable. Participants tended to perform videogame training in short sessions with frequent pauses, concentrated in the evening hours before the expected bedtime for each age group. Evenings were likely to be when participants were the most fatigued, but also when they had the most free time available to play the treatment videogame. Children likely had shorter attention spans compared to older participants, leading to even shorter game-playing sessions and more frequent pauses. This pattern of adherence in a home-based treatment trial on a portable device, which found a negative outcome, stands in contrast to previous successful adult studies in laboratory[8, 9] settings where training was usually completed in hour-long blocks during day-time office hours under supervised conditions with minimal distractions.

A large portion of our participants over-reported treatment adherence in their training diary despite knowing the game software was objectively monitoring their adherence. The worst cases of over-reporting tended to occur in younger children, which may reflect difficulties in relying on parents or caregivers to record the diary for the child. Our results confirm experience from other clinical trials[3, 13, 14] that objective recording of treatment adherence is essential in both research and clinical settings. Relying on subjective report alone carries a high risk of over-estimating treatment adherence, leading to under-estimation of treatment efficacy.

Treatment adherence gradually fell over the 6-week videogame treatment period, mainly due to participants missing days of training entirely rather than playing less on each day. This pattern is similar to treatment adherence observed in electronically-monitored patching therapy for amblyopia,[3] though we observed a less dramatic decline likely due to the shorter duration of binocular treatment (42 days) compared to typical occlusion therapy (median 99 days, IQR 72 days). Many of our participants mentioned boredom with Tetris by the 3 weeks follow-up visit, and reported falling motivation to maintain adherence for the second half of the 6-week treatment period. This suggests that future binocular treatments should improve engagement using age-appropriate game mechanics[16], offer more gameplay variety[21] and/or include other visual content such as movies or cartoons[6, 22] to maintain active engagement and motivation for the entire duration of treatment.

Frequent game pauses seen in our data confirm anecdotal reports that participants may have been inattentive or multitasking (for example some may have been concurrently watching television) and thus not continuously viewing the binocular stimuli presented in the active game. Binocular therapies are hypothesised to treat amblyopia by providing prolonged periods of binocularly-balanced visual stimulation.[4] Multiple brief exposures with frequent disengagement, as seemed to occur in the current study, may carry a reduced treatment effect compared to the long duration exposures that characterise laboratory-based training. However, the effects of interrupted binocular stimulation have so far not been specifically investigated.

Potential disengagement and dose-discontinuity complicates dose-response calculations for binocular amblyopia treatment, which is currently based solely on cumulative gameplay duration.[13-16] Future studies would benefit from gaze tracking to precisely monitor exposure patterns to dichoptic stimuli. Gaze monitoring can also be used to prompt the patient if they become distracted or looked away. A currently ongoing clinical trial of binocular amblyopia treatment has included gaze tracking for monitoring treatment adherence[23], but data are not yet available.

It may be that for treatments based on visual-stimulation to be successful, detailed instructions need to be given to patients (and parents/caregivers where relevant) specifying the optimal session length and emphasising the need to continuously view the display for maximum effect. However, continuous viewing must be enforced in a friendly manner to be acceptable to the user, and longer training sessions need to be balanced against the fact that they are less convenient to schedule within the patient’s daily routine. Inconvenient or unpleasant treatments will hinder overall treatment adherence, which can also lead to reduced effectiveness. This balance between training intensity and practicality is an important design consideration for all home-based treatments, and the optimal balance will depend on many factors including patient age, ability, lifestyle factors (e.g. school, work, and/or family responsibilities), the treatment delivery method (e.g. portable versus not portable), and, importantly, the true impact of disrupted play on treatment outcomes, which may vary depending on the condition being treated and the mechanisms underlying the treatment effects. Traditional vision therapy exercises for vergence disorders, for example, are often prescribed with a “little but often” daily regimen to minimise ocular fatigue. But this may not be suitable for all training-type treatments.

## Conclusions

Adherence to home-based videogame treatment for amblyopia was often less than prescribed and frequently over-reported. Self-administered at-home treatment includes frequent interruptions, particularly in children, which may reduce effectiveness of treatment methods that rely on continuous stimulation. Objective monitoring, including gaze tracking for attentive eye-to-screen durations, is required to assess the true doses of dichoptic stimulation received and examine potential dose-response relationships of visual treatments.

## Data Availability

The main visual outcomes of the optical treatment and videogame treatment phases of the BRAVO clinical trial are available in the following two publications: 1) Optical treatment phase outcomes: Gao, T.Y., et al., Optical treatment of amblyopia in older children and adults is essential prior to enrolment in a clinical trial. Ophthalmic Physiol Opt, 2018. 36(2): p. 129-143.
2) Videogame treatment outcomes: Gao, T.Y., et al., Effectiveness of a Binocular Video Game vs Placebo Video Game for Improving Visual Functions in Older Children, Teenagers and Adults with Amblyopia: A Randomized Clinical Trial. JAMA Ophthalmol, 2018. 136(2): p. 172-181.
De-identified datasets may be supplied on reasonable request, following approval of the trial Steering Committee. Please contact the corresponding author Dr Tina Gao or Prof Benjamin Thompson.

## Acknowledgements

**Disclosure Statement:** Authors Benjamin Thompson and Robert F. Hess are named inventors on two patents (US 12528934 and US 8006372 B2) related to the contrast-balancing binocular videogame treatment used in the BRAVO clinical trial. Robert F. Hess is a scientific advisor to Amblyotech, a company that licenses the patents.

All other authors do not have conflicts of interest or competing financial interests.

**Study Funding:** The BRAVO clinical trial was supported by research project grants from the Health Research Council of New Zealand (reference 13/169) and the Hong Kong Health and Medical Research Fund (reference 11122991). The Centre for Eye Research Australia receives Operational Infrastructure Support from the Victorian Government.

The funding bodies had no role in the design and conduct of the study; collection, management, analysis, and interpretation of data; preparation, review, or approval of the manuscript; or the decision to submit the manuscript for publication.

